# Does sugar control arrest complications in type 2 diabetes? Examining rigor in statistical and causal inference in clinical trials

**DOI:** 10.1101/2022.08.02.22278347

**Authors:** Akanksha Ojha, Harshada Vidwans, Milind Watve

## Abstract

In contrast with type 1 diabetes mellitus (T1DM), in type 2 (T2DM) the success of intensive glucose normalization in arresting diabetic complications is marginal and inconsistent across multiple clinical trials. However, glucose regulation still largely remains the main target of treatment for T2DM in clinical practice. We examine the scientific rigor behind the design, conduct and inferences of 6 major clinical trials targeting glucose normalization and following up for diabetic complications and mortality. We find and discuss multiple flaws in reporting the results, their statistical treatment and clinically useful recommendations. The most serious flaw is the inability to recognize the limitations of statistical inferences when multiple comparisons are involved. Further we show using simulations that when different outcomes are not independent of each other, significance gets overestimated. We also suggested alternative ways to assess the effect of antihyperglycemic treatment, if any. Using more sound and elaborate statistical methods and inferential logic we find no support to the prevalent belief that intensive glucose normalization has any benefit in terms of reducing the frequency of any of the complications. Furthermore, alternative interpretations of the results have not been considered and evaluated in any of the clinical trials or their meta-analysis so far. Because of failure to show consistent significant benefit across multiple trials, we should now treat the hypothesis that glucose normalization prevents complications in T2DM as decisively falsified. This necessitates rethinking about some of the fundamental beliefs about the pathophysiology of diabetic complications and facilitate novel alternative lines of research.

---

> “Absolute proof that good (glycemic) control can retard or prevent the development of complications has not yet been obtained, but the assumption is accepted by most diabetologists and indeed is almost an article of faith in the current approach to achieve the best possible diabetic control” Geoffrey Gill (1991).

Diabetes was first recognized by the appearance of sugar in the urine a long time ago. Later increased glucose in blood became known as the marker of diabetes. Insulin was discovered about 100 years ago and our thinking in diabetes has mostly revolved around the two molecules although now over a hundred molecular, cellular and neuronal signals are known to be altered in diabetes (Watve, 2013). Network models trying to integrate all these signals have raised doubts about the central and causal role of glucose and insulin in the pathophysiology of diabetes (Kulkarni et al., 2017), but the mainstream clinical thinking and action continues to revolve around glucose and insulin. The differentiation of type 1(T1DM) and type 2 diabetes mellitus (T2DM) gradually became clear over a few decades by the mid 20^th^ century (Colman et al., 1999; Dana, 1954; Harris, 1988; Himsworth, 2013; Null et al., 1973; Yki-Järvinen et al., 1986). Insulin was highly successful not only in regulating glucose but also in arresting complications in T1DM (Fullerton et al., 2014; Nathan, 2014; The Diabetes Control and Complications Trial Research Group, 1993). Owing to the perception that T1DM and T2DM differ only in the cause of glucose dysregulation and that the downstream pathophysiology originating from chronic hyperglycemia is almost identical in the two types, the success of glucose regulation observed in T1DM was expected to work for T2DM as well.

However, from early clinical trials of glucose regulation in type 2 diabetes, it was clear that glucose regulation in T2DM was not as effective in arresting diabetic complications as in T1DM (Huang et al., 2001; Skyler, 1978). The debatable issue has been whether it is effective at all (Boussageon et al., 2017). The ultimate goal of drugs used in treatment of T2DM should be prevention of complications, but the immediate surrogate goal is believed to be to normalizing glucose. There are two common assumptions behind treatments targeting glucose normalization (i) increased glucose is causal to diabetic complications and (ii) normalizing glucose can reduce the frequency of complications with statistical and clinical significance. The first major blow to this set of assumptions came from the results of the University Group Diabetes Program (UGDP) in which glucose normalization failed to decrease complications and mortality, instead it increased in some of the treatment arms (Knatterud 1978). UGDP came under some criticism and controversy and a number of other clinical trials followed in subsequent decades that used different study designs, sample sizes, drugs used and outcome measures examined. While some of them claimed that the treatment significantly reduced certain adverse outcomes, others had a perplexing finding of increased cardiovascular and all cause mortality in the intensively controlled group (Ismail-Beigi et al., 2010; The NICE-SUGAR Study Investigators, 2009). In order to address the contradicting results, particularly in the macrovascular outcomes, and raise a coherent picture, a collaboration was established between four major trials namely UKPDS, ACCORD, ADVANCE and VADT. A meta-analysis of the four trials concluded that a “modest decrease” in macrovascular events was achieved by intensive glucose regulation (Turnbull et al., 2009).

However, a number of issues remain unaddressed regarding the scientific rigor in the designs of different trials, the statistical treatments used and inferences drawn from the results. We discuss here the various possible traps and biases in the studies, possible alternative interpretations of the results and ultimately the scientific soundness of the current set of assumptions in the treatment of type 2 diabetes. We use a comprehensive analysis of large scale institutional, multicentre, multi-drug trials of glucose normalization with long term follow up, broadening the scope of the Turnbull et al (2009) meta-analysis, re-examining the design, statistical analysis and inference related issues and clinical usefulness.

The necessity of being sceptical about published randomized clinical trials (RCT) and cross examination of its validity has become a critical issue over the last decade or two (Chow et al., 2021; Heneghan et al., 2017; Herrera-Perez et al., 2019; Ioannidis, 2016). There have been multiple attempts to bring in reforms and rigor in RCTs. One of the measures is to make registration of the RCT mandatory before recruiting participants. Issues addressed by mandatory registration include failure to report, partial reporting, hiding inconvenient results, changing the norms of studies, issues of data transparency etc (James et al., 2015; Ramsberg & Platt, 2018; Thabane et al., 2015; Tse et al., 2009; Zarin & Keselman, 2007). However, in spite of the increasing awareness and attempts to improve clinical trial designs, many issues still remain unaddressed. Furthermore, many of the clinical trials for glucose normalization in T2DM began prior to the recent measures to refine clinical trial designs. Therefore it is essential to specifically examine the possible biases in glucose regulation trials for T2DM and their effects on the robustness of inferences and clinical usefulness.

## Selection of studies and inclusion exclusion criteria

We focus on both qualitative and quantitative issues in this article. For quantitative analysis in this study we selected institutional clinical trials having a minimum of 100 individuals per group, minimum follow up of 3 years and reporting rate ratio (RR), odds ratio (OR) or hazards ratio (HR) (or data sufficient to calculate these indices) for multiple macro and microvascular events and mortality. We excluded those that reported only surrogate outcomes. We excluded trials conducted by pharmaceutical sector companies during drug development. The trials selected by these inclusion-exclusion criteria comprise University Group Diabetes Program (UGDP), the UK prospective diabetes study (UKPDS), the Veterans Affairs Diabetes Trial (VADT), the Action in Diabetes and Vascular Disease: Preterax and Diamicron Modified Release Controlled Evaluation (ADVANCE), the Action to Control Cardiovascular Risk in Diabetes (ACCORD) and the diabetes prevention programme (DPP).

For analysis we take all the comparisons between a treatment or intensive treatment group with a control group without the specified treatment regime with or without blinding/placebo. We exclude comparison between drugs. We also exclude trial arms with specific drugs whose use was discontinued, e.g. tolbutamide in UGDP or troglitazone in DPP.

The end points/outcomes included in analysis were those that have at least 10 events in at least one of the groups being compared. We considered all single end points and aggregated outcomes reported in tables, figures as well as in the text. It is not clear why some results were reported in tables while some were reported in the text only. But it was thought necessary to evaluate using everything that is reported. We excluded any subgroup analysis. We also excluded any associative analysis that tries to relate the glycemic status with the incidence of complications by ignoring the allocation to trial regimens such as in (Hayes et al., (2013); or Stratton, et al (2000).

Since the trial designs, characteristics of patient groups, definitions of outcomes used vary across studies, and since a meta-analysis approach has been used before, we do not use a meta-analysis approach but instead look at the results as reported an a collective and critical way with a focus on issues that remain unaddressed or inadequately treated in earlier literature.

We observe that the main issues unaddressed or inadequately addressed across the selected trials are as follows.

1. The total number of individually significant outcomes: Summing up over the 6 clinical trials (UGDP, UKPDS, VADT, ACCORD, ADVANCE and DPP) out of the total 341 outcomes compared between intensive and conventional control groups, in 38 pairs the frequency of adverse outcomes is individually significantly lower than the control and in 26 it is individually significantly higher. However there are repetitions and interdependence in them, for example all cause mortality includes cardiovascular mortality and thereby if the latter is significantly different, the chances of the former being significant increase. Eliminating such obvious duplications, significant reduction is seen for 17 and significant increase for 13 adverse outcomes summed up over all the 6 trials. By this simple summation, the beneficial outcomes cannot be said to be significantly greater than the harmful outcomes. If soft, subjective and surrogate end points are removed, no difference in the number of beneficial and harmful results remains.
2. Appropriate use of significance level when multiple comparisons are made: When multiple statistical comparisons are made, it is possible that some of them turn out to be “significant” by chance alone. This is a well-known statistical problem and the suggested solution is to decrease the significance level in proportion to the number of statistical tests used (Narum, 2006, Rice et al 2008). None of the clinical trials have used any of the suggested corrections for significance level used. Turnbull et al (2009) recognized this problem but did not attempt any solution. The total number of outcome measures reported in these trials is large. If the Bonferroni principle or any other correction for significance level is applied, none of the effects of treatment turn out to be significant. Since the Benferroni correction is too conservative, we look at how many outcomes are expected to be significant at 0.05 level by chance alone and whether the number of individually significant results observed in the trials are significantly greater than the expected. However, when multiple statistical tests are conducted, how many will turn out to be individually significant by chance alone is not an easy question to answer when the different outcomes are not independent. There are two types of dependencies in the glucose normalization trial data. One is that of repetition in aggregate end points as mentioned above. The other is that of having common pathways/mechanisms behind many outcomes. If the common pathway happens to differ between the two groups by chance, a number of outcomes may appear to be significant together. This problem has not been addressed in the statistical treatment of clinical trials with multiple outcome measures so far. We address this problem using simulations (see supplementary material for details of the simulation model). The simulations generate the control and treated group and the outcome in each individual in the group is generated by appropriate randomization. The ORs resulting from these simulations are studied. In the baseline simulations where treatment effect is assumed to be zero and each outcome is assumed to be independent and have the same parameters, the ORs are distributed with a mode around unity. Being ratios, the distribution is positively skewed as expected and the ORs at both the tail ends are individually significant. However, in reality the incidence of every outcome is substantially different, for example, in the ADVANCE trial visual impairment was seen in 54 % patients but dementia in only 0.9%. When differences in incidences are incorporated in the simulations, not only the tails but ORs with intermediate departure from unity on either side are more likely to be significant. This is because at low incidence, a greater departure from unity is possible by chance alone, but significance is difficult owing to the smaller number of cases. On the other hand with larger incidence, moderate departures from unity also turn out to be individually significant. This pattern matches with the clinical trials data (figure 1).

**Figure 1:**
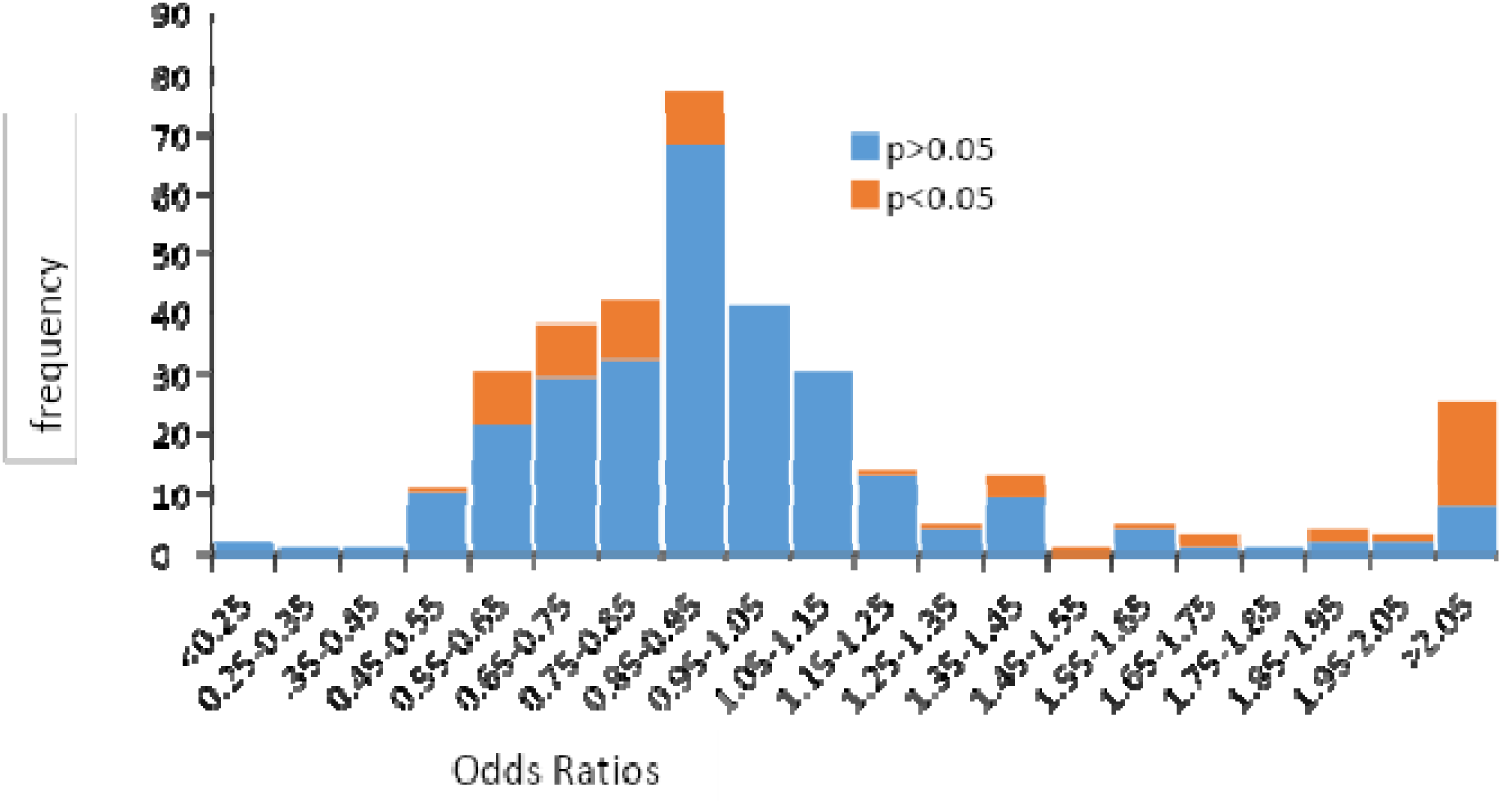
The frequency distribution of odds ratios for all outcomes reported in the six clinical trials. The mode and median have shifted slightly to the left from unity, but this difference cannot be said to be statistically significant. Individually significant statistical deviations are on both sides. The geometric mean of the population is close to one (1.01) indicating near symmetry in magnitude. The simulations can incorporate interdependence in different ways. By the classical assumption hyperglycemia is the common trigger behind all pathophysiology of diabetic complications. If this assumption is true, all outcomes should be interdependent. Alternatively it is also possible that some clusters of outcomes are interdependent but independent of other clusters. This assumption is relevant to T2DM since the factors involved in microvascular and macrovascular complications can be different. Further in some tissues capillary density appears to reduce where as there is hyper-angiogenesis in some other complications. Simulations show that by either modes of dependence, in a large proportion of simulation runs the proportion of outcomes individually significant can be substantially greater than the expected 5%. Therefore having greater than 5% outcomes individually significant cannot be taken to reflect the effect of treatment. Since we do not have empirical estimates of the extent of interdependence, we cannot state how many outcomes can turn out to be individually significant by chance alone. A possible solution to the problem is that instead of depending upon individual level significance, we look at the distribution of all ORs. If there is a favourable treatment effect on the common mechanism itself, as is assumed by the classical theory of T2DM, then the entire distribution should shift to the left. The significance of this shift can by judged by multiple simulation runs. We observe that when pooled over all the six trials, the distribution of ORs has a mode and median shifted to the left by 10 % and 7.3 % respectively (figure 1). By this indication, intensive glucose control may be said to have a benefit of 7 to 10 % which is compatible with the estimate of 9 % obtained by Turnbull et al (2009). This contrasts with the distribution of ORs in T1DM where almost the entire distribution of ORs lies to the left of 1 (Fullerton et al., 2014; Nathan, 2014; The DCCT Research Group, 1993). Similarly for exercise intervention for T2DM across studies, almost all ORs are less than 1 (Sluik et al., 2012). The distribution of ORs in the T2DM trials has only a marginal leftward shift. Whether this shift is significant or not is questionable. In simulations comparing 350 outcomes, comparable to the pooled data of all 6 trials, 10% or greater shift in the mode and the median was observed in about 16 % of simulation runs (see Supplementary information). Therefore the shift observed in the pooled 6 trial data may not be considered significant. If we assume that some clusters of outcomes have some common underlying pathway and are therefore interdependent, it is seen that a greater than expected proportion of outcomes can turn out to be individually significant by chance alone in a substantial proportion of the simulation runs. However, they may be symmetrically or asymmetrically distributed towards the two tails. Two or more simulation runs can result in significant heterogeneity among them although they are run at the same parameters. Since we do not have empirical data on the extent of interdependence, simulations cannot be used to make quantitative predictions. But they conclusively show that if the assumption of independence of chance acting on every outcome is violated, the number of false positive significance increases substantially. Unless this effect is accounted for, we cannot conclude that the reduction in some of the outcome measures claimed in the treatment groups is indeed significant and not a false positive. It is equally likely that the significant rise in mortality seen in certain trials such as tolbutamide arm in UGDP, the metformin plus sulfonylurea group in UKPDS or in the rosiglitazone trial (RECORD)(Home et al., 2009) also may be chance alone. If this is true, other inferences such as glucose normalizing treatment being more effective for microvascular outcomes than for macrovascular ones is equally unfounded. Further the observed heterogeneity between trials (Turnbull et al., 2009) is also possible by chance alone. In short since we know that certain common mechanisms are involved in the pathophysiology of diabetic complications, we should expect a large number of false positive results as well as heterogeneity between trials. On this background, the observed individual outcomes of the trials are marginal and therefore any inferences cannot be confidently made. The shift in the mean, mode or median of the distribution is also not significant and therefore the hypothesis that hyperglycemia is the common pathophysiological mechanism of all complications and anti-hyperglycemia treatment can arrest complications is not supported.
3. Placebo effects: Having a double blinded placebo controlled design is a standard norm for almost all clinical trials, unless a specific context makes blinding impossible. While ACCORD, ADVANCE, VADT, UGDP and DPP have some kind of blinding or placebo control, whereas UKPDS is an open label trial. Plotting the distribution of ORs in the trials with and without placebo reveals that the shift in mean and median to the left is observed for UKPDS alone which does not have a placebo control (figure 2). The distribution of ORs in the pooled data of trials with placebo have a mode at 1 and do not show a left shifted distribution of ORs. This raises the possibility that the apparent benefit of treatment claimed in UKPDS may only be a placebo effect. DPP allows comparison of a placebo group with a no medication group within a single trial. There the difference is not statistically significant but is of a comparable order (Lee et al., 2021). Some other studies have also demonstrated detectable placebo effects in diabetes treatment (de Wit et al., 2016; Sievenpiper et al., 2007).

**Figure 2:**
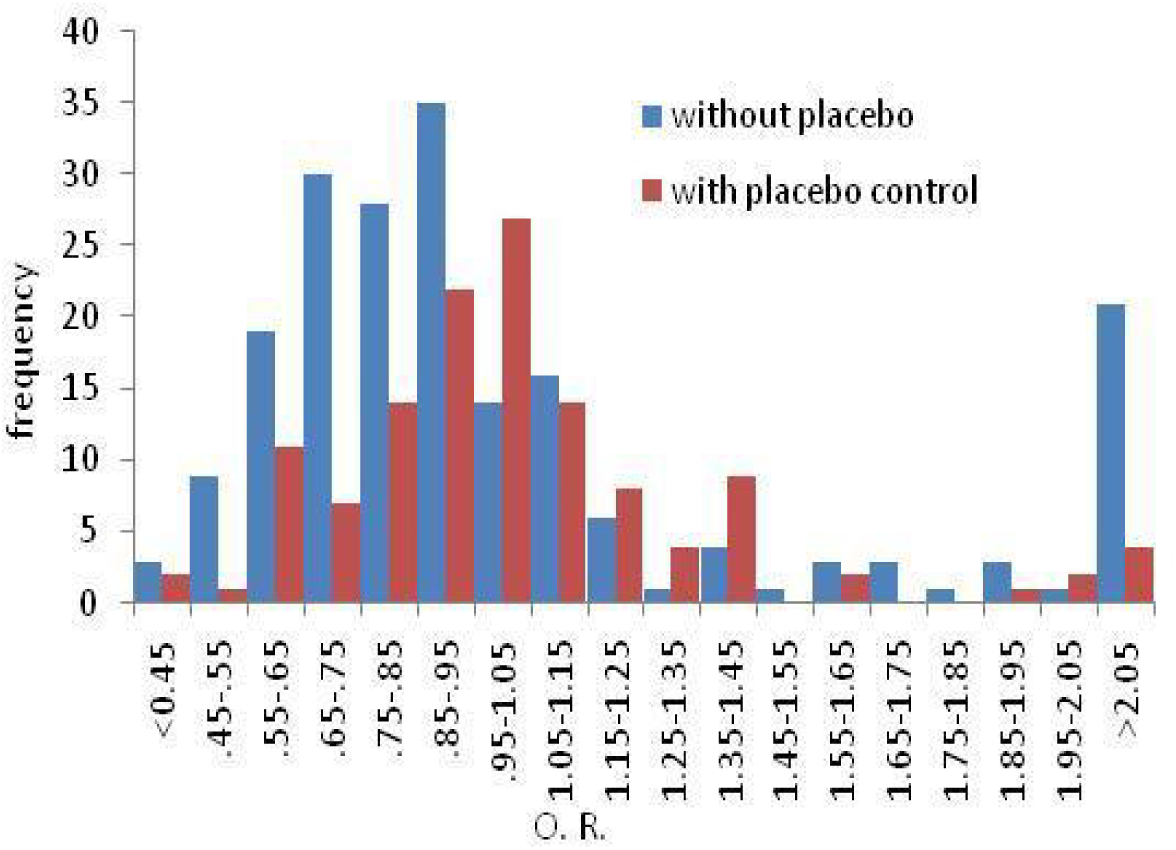
The distribution of ORs in trials with and without placebo control. There is a leftward shift in the distribution without placebo control. The marginal leftward shift seen in the total (figure 1) can therefore be suspected to be a placebo effect. In clinical trials with surrogate markers, placebo effect can potentially operate at two distinct levels. One is in receiving the drug versus the blank. The second possible level of a placebo effect i.e. knowing that my blood sugar is under better control can potentially have positive psychosomatic effects. If a reduction in the frequency of complications or mortality is observed, it may be potentially explained by the second level placebo effect. So far, to the best of our knowledge, no trial has attempted to address the potential second level placebo. Unless this possibility is seriously considered, tested using appropriate controls and rejected, the marginal beneficial effects observed cannot be confidently claimed to be due to better glucose regulation.
4. Even if we assume that the marginal benefits observed in some of the outcomes are not by chance, it does not show that they are a result of glucose regulation. Across as well as within trials, the difference in HbA1c achieved and the relative success in preventing complications do not always correlate well. Within UKPDS, the sulfonylurea and insulin arms had HbA1c difference of 0.9 but a significant difference was seen only in the soft endpoint of need for retinal photocoagulation (UK Prospective Diabetes Study (UKPDS) Group, 1998a). In the post trial follow up the difference in HbA1c had vanished but a greater benefit of intensive control was apparent (Holman et al., 2008). Unlike UKPDS, a post trial long term follow up in VADT and DPP did not show any long term benefits (Reaven 2019, Lee et al 2021) demonstrating the lack of consistency across trials. In the overweight patients group of UKPDS, the advantage obtained by metformin over sulphonylurea and insulin was not explicable by better glycemic control (UKPDS group 1998b). Across trials ACCORD and ADVANCE achieved the largest HbA1c difference, but they do not exhibit the greatest success in arresting complications. Other studies (e.g. Stratton et al 2000) show an association of glycemic status with the risk of complications, but these studies cannot be used to make a causal inference. It is possible that for individuals with a more serious underlying pathology, the chances of complications are higher as well as glycemic control is more difficult. This results in a correlation but does not imply that reducing glycemia would result in risk reduction. Many of the drugs used in treatment of T2DM have multiple effects in the body independent of glucose regulation. Insulin is known to have multiple functions in the body ranging from amino acid metabolism, cell division to cognitive function and ovulation regulation (Shemesh et al., 2012; Strachan, 2003). Metformin is shown to affect endothelial function directly (Diamanti-Kandarakis et al., 2005; Heidari et al., 2019). The beneficial effects of SGLT2 inhibitors are seen in diabetic and non-diabetics alike (DeFronzo et al., 2019; Ferrannini & DeFronzo, 2015). Therefore even if any beneficial effects of any of the drugs in diabetes treatment are seen, it cannot be stated that the benefits are because of improved glucose regulation. Even in T1DM whether the observed efficacy of insulin treatment is because of glucose regulation or because of the multiple other functions of insulin has not been clearly addressed and resolved.
5. The magnitude of difference: Although the number of outcomes with individually significant beneficial effects of treatment are marginally greater than the number of outcomes with increased frequency, the frequency of harmful effects is substantially greater. While the individually significant reductions in ORs range between 0.5 and 0.92, the increase ranges between 1.2 and 13.1. The harmful effects of intensive treatment not only include major hypoglycaemic events and other drug related symptoms, but also cardiovascular mortality, cancer and all cause mortality in different trials. By the properties of ratios, the distribution of OR is bound to be positively skewed. The geometric mean of all ORs is close to one (1.01) indicating that the net odds ratios do not deviate from unity. Therefore it cannot be stated that the benefits, if any, of intensive glucose regulation outweigh its harmful effects. Even if, for the time being, we ignore the harmful effects and focus on the beneficial ones alone and assume that the claimed significance was real, whether the benefits are clinically meaningful is a crucial question. Although in public health literature, the emphasis is on ratio based indices such as OR, RR and HR, common people seem to go by absolute risk reduction (ARR). In an experiment, respondents including the ones having undergone training in public health statistics were observed to judge their own risk by probability difference rather than by probability ratio (Vidwans et al 2021). In all glucose regulation trials for T2DM, the ARR is too small to be clinically meaningful. For example, in ADVANCE trial over a 5 year follow up, the incidence of combined major microvascular and macrovascular events was 20 % in the control group and 18.1 % in the treatment group. Although by ratio based indices this is about 10 % reduction, by difference based indices it is only 1.9 %. The number needed to treat (NNT) for preventing diabetic complications in any of the trials is 20 and above even after ignoring the increased frequency of some adverse events. Since different trials have different follow up periods, and the effect appears to be more or less linear consistently over time, we can express it as NTNT, i.e. number-time (person-years) of treatment needed for preventing one complication (Laupacis et al., 1988). NTNT is fairly consistently distributed across trials and averages to about 250. That is if 25 diabetics are treated targeting glucose regulation for 10 years, one complication in one of the 25 patients may be prevented. This is quite a low success rate. Whether at this success rate it is worth undergoing treatment for glucose control, that has a considerable frequency of adverse events is a subjective decision which should be left to the patient after having informed about NTNT in a simple language. Failure to inform the patient about the efficacy of treatment seen in clinical trials potentially amounts to a violation of human rights.

Apart from the trials included here, there are many more that converge on the same inference. These trials did not fit into our inclusion exclusion criteria but their inferences converge with our analysis. The NICE-sugar study revealed that under critically ill patients tight sugar control resulted in higher mortality (The NICE-SUGAR Study Investigators, 2009). Huang et al (2001) meta-analyzed 5 glucose regulation trials for their effect on cardiovascular disease. Apart from UGDP and UKPDS they included VACSDM, Kumamoto and DIGAMI trials and the meta-analysis yielded no effect of glucose lowering on cardiovascular disease.

In short, owing to multiple flaws in the design, statistical analysis and inferential logic in the clinical trials of glucose regulation in T2DM, at present we cannot state with scientific rigor that glucose regulation has any clinical benefit in T2DM, in contrast with T1DM.

This brings into question the classical assumption that T1DM and T2DM differ only in what goes wrong in glucose regulation and all downstream effects of hyperglycemia are similar. It is possible that the two are fundamentally different and converge only on one of the symptoms that is hyperglycemia. Hyperglycemia may not be central to the pathophysiology of the two in a similar way. Therefore the target of treatment of the two needs to be different.

### Evidence for confirmation bias

Although the difference between the response of T1DM and T2DM to treatment targeting glucose regulation is evident for quite a few decades, there is a deep rooted reluctance to accept the evidence and refute glucose lowering as the main line of treatment in T2DM. We argue that this is because of confirmation bias which is evident in multiple ways in the analysis and presentation of data in the publications resulting from the clinical trials.

a. Reporting positive and negative outcomes with different statistical treatment: At times, for reporting two different results in the same paper, two different reporting formats are used without justifying the difference. Quoting verbatim from the reported UKPDS results (UKPDS Group, 1998a),

> “Compared with the conventional group, the risk in the intensive group was 12% lower (95% CI 1–21, p=0· 029) for any diabetes-related endpoint; 10% lower (–11 to 27, p=0 · 34) for any diabetes-related death; and 6% lower (–10 to 20, p=0 · 44) for all-cause mortality.” In the following paragraph, the risk of major hypoglycemic events is reported as,

> “The rates of major hypoglycaemic episodes per year were 0 · 7% with conventional treatment, 1 · 0% with chlorpropamide, 1 · 4% with glibenclamide, and 1 · 8% with insulin.” It is strange that the risk reduction is given as percent reduction, but the incidence of hypoglycemic events reported as absolute frequencies. If the risk of hypoglycemic events is also expressed as percentage, with chloropropamide it is 43% higher, with glibenclamide it is 100% higher and with insulin it is 170 % higher. Effectively the increase in risk of major hypoglycaemic event is up to an order of magnitude greater than the maximum reduction seen in any of the complications in the trial. However, since the reporting format is different, a common reader is more likely to attach greater importance to percentage figures and attach smaller importance to hypoglycaemic events since the numbers reported are fractional. The tradition of reporting favourable and unfavourable results using different statistical expressions is continued in many other clinical trials of T2DM. Another way of differentially reporting favourable and unfavourable results is including them in the figures or tables versus making a passing mention in the text. In UKPDS as well as in DPP many outcomes are mentioned in the text but not included in the tables/figures. The text sometimes has incomplete information to calculate OR, confidence intervals or statistical significance. Interestingly not a single outcome reported in the text shows significant favourable effect of treatment. There is no justification offered why these were not included in the tables. In DPP, for example, medication related gastrointestinal symptoms are mentioned in the text but not included in the tables. Participants that were unable to continue metformin due to adverse reaction are mentioned in the text, but these adverse events do not seem to be included in the analysis (The DPP research group 2012).
b. Cherry picking: The UKPDS results (UKPDS group 1998a) say that the reduction in all diabetes related end points was 12% and statistically significant. However, this does not include major hypoglycemic events. If major hypoglycemic events are pooled with other adverse events, no different between control and treated group remains w.r.t. all outcomes. The primary reports of UKPDS enlist a large number of single point and aggregate outcome measures, but the post-trial follow up reports only seven aggregate outcomes. There is no way to know whether only the ones showing favourable results were selectively reported.
c. Changing targets: The UKPDS study first decided to consider at least 40% reduction in mortality or morbidity in the intensive treatment group in order to be clinically significant. This benchmark was reduced to 15 % subsequently. But UKPDS has been criticised for not maintaining even the lowered standard and later declaring a 12% reduction as significant (Ewart et al., 2001).
d. Recognizing but Failing to correct flaws: The meta-analysis of 4 trials (Turnbull et al., 2009) recognizes the flaw that the necessary adjustment of significance level for multiple statistical comparisons has not been made, but does not try to apply any correction for it. The published reports of individual trials do not even acknowledge this serious flaw, which is quite well known in the field of statistics.
e. Manipulating readers’ perspective: Most readers stop after reading the title and abstract. Therefore selecting what to write in the title and abstract is an effective way to manipulate readers’ perspective. For example the meta-analysis by Turnbull et al (2009) acknowledges that since they have not made correction for multiple comparisons, their analysis should be treated as tentative and exploratory, this admission does not figure in the abstract and therefore most readers after reading only the abstract are likely to take their results as robust and conclusive.

The most parsimonious explanation of all the reporting anomalies together is that it comes out of inadvertent or intentional confirmation bias and the burden of history. Since the substantial benefit of insulin treatment in T1DM was clearly shown, it was expected that it should work for T2DM as well. Since the assumed pathophysiology was similar, the expectation was strengthened. On this background, it would be natural to be reluctant to accept counterintuitive results. Confirmation bias comes out of human nature and may not be treated as a crime, but there needs to be a continued attempt to overcome it and face the reality above prior beliefs and prejudices. Going by the results of clinical trials, considering all alternative possible explanations of the results it needs to be clearly recognized that there is no conclusive evidence so far that glucose normalization reduces diabetic complications in the context of T2DM. The remark by Gill (1991) quoted at the top of the article is applicable even after over 30 years of research. The failure of multiple attempts to support the hypothesis that glucose regulation can arrest complications in type 2 diabetes should be taken as decisive and the target of treating T2DM changed accordingly. Although this is being increasingly recognized (Boussageon et al., 2017) the recognition is still fragmentary. The Lancet commission on diabetes (Chan et al., 2020) clearly recommends “access to insulin, patient education, and tools for monitoring blood glucose” only in the context of T1DM now and for T2DM, instead of emphasizing on glucose regulation, it recommends addressing “diverse environmental, behavioural, and socioeconomic causes” and “sustained reduction of common cardiometabolic risk factors”. However, the changing stance in literature has not yet reflected sufficiently in clinical practice. This needs active efforts to simultaneously educate practitioners as well as common man. Treatment of T2DM is certainly going to be a case of “medical reversal” i.e. a long standing treatment recommendation based on a belief is completely withdrawn based on evidence (Herrera-Perez et al 2019). Facilitating the process of medical reversal along with research on effective alternatives will be critical for arresting one of the greatest health concern of the present and future.

## Supporting information

We address this problem using simulations (see supplementary material for details of the simulation model).

## Data Availability

All data produced in the present study are available upon reasonable request to the authors

## Ackowledgements

This study benefitted from an ongoing systematic review of all clinical trials in T2DM conducted by Shubhankar Kulkarni, Pramod Patil, Vibha Bapat, Hrishikesh Chunekar, and Varada Mengale. We thank Vikrant Patil for writing the code for the simulation program.

## Notes

### Competing Interest Statement

The authors have declared no competing interest.

### Funding Statement

This study did not receive any funding

