## Supplementary material for "Does sugar control arrest complications in type 2 diabetes? Examining rigor in statistical and causal inference in clinical trials": We address this problem using simulations (see supplementary material for details of the simulation model).

Akanksha Ojha^1,2^

Harshada Vidwans^3^

Milind Watve^1,4^ *

1. Deenanath Mangeshkar Hospital and Research Centre, Pune, India

2. Indian Institute of Science Education and Research, Pune, India

3. Independent Researcher, B 104, Rutuparna Society, Baner, Pune 411045, India

4. Independent Researcher, E-1-8, Girija Shankar Vihar, Karvenagar, Pune 411052, India.

Abstract:

In contrast with type 1 diabetes mellitus (T1DM), in type 2 (T2DM) the success of intensive glucose normalization in arresting diabetic complications is marginal and inconsistent across multiple clinical trials. However, glucose regulation still largely remains the main target of treatment for T2DM in clinical practice. We examine the scientific rigor behind the design, conduct and inferences of 6 major clinical trials targeting glucose normalization and following up for diabetic complications and mortality. We find and discuss multiple flaws in reporting the results, their statistical treatment and clinically useful recommendations. The most serious flaw is the inability to recognize the limitations of statistical inferences when multiple comparisons are involved. Further we show using simulations that when different outcomes are not independent of each other, significance gets overestimated. We also suggested alternative ways to assess the effect of antihyperglycemic treatment, if any. Using more sound and elaborate statistical methods and inferential logic we find no support to the prevalent belief that intensive glucose normalization has any benefit in terms of reducing the frequency of any of the complications. Furthermore, alternative interpretations of the results have not been considered and evaluated in any of the clinical trials or their meta-analysis so far. Because of failure to show consistent significant benefit across multiple trials, we should now treat the hypothesis that glucose normalization prevents complications in T2DM as decisively falsified. This necessitates rethinking about some of the fundamental beliefs about the pathophysiology of diabetic complications and facilitate novel alternative lines of research.

Simulations to see the effect of interdependence of outcomes in a multi-outcome clinical trial

Purpose: In a clinical trial there is typically a group undergoing treatment along with a control group. The groups are randomized to begin with and preferably double blinded, i.e. neither the participants nor the persons monitoring the outcomes know who belongs to the treated or the control group.

The problem that we are addressing here is that if in a single trial multiple end points or outcomes are being tested, then in some of them the difference between the control and treated group may turn out to be “significant” by chance alone. When we use 0.05 as the conventional significance cut-off, about 5 % of the differences are likely to turn significant by chance alone. This significance does not mean that the treatment is effective. In some clinical trials of glucose normalization in type 2 diabetes, up to 121 outcome measures were recorded and we can expect that some of them may turn out to be significant even when the treatment has no effect. This is a well recognized problem and the solution suggested is to lower the significance cut off probability when performing multiple statistical procedures.

However all the possible solutions to the multiple testing problems are based on the assumption that chance affects each test independently. That may not be true in clinical trials. Particularly in diabetes one or more common pathways are believed to be responsible for many outcomes. If one of the common mechanisms/pathways happens to be coincidently favorable/unfavorable for many participants in the treatment group, suddenly a large number of outcomes may turn out to be significant. In addition, in the glucose regulation based clinical trials of T2DM, single end points as well as aggregates are considered. Since the aggregate end points are summations of many single end points, there is dependence in them. In situations where the assumption of independence of chance element for every outcome is violated, how should one treat it statistically is the question.

We use simulations to examine at a given level of dependence how the resultant statistic is distributed, and whether based on these distributions we can make any statements about the probability of false positive significance. This principle can then be applied to interpret the results of clinical trials recording multiple outcomes.

The model:

A baseline model assuming independence: We generate two randomized groups of participants with the same size n. For each outcome there is an incidence *i* (always <1 ) which varies within a range. The treatment effect, if any, is *t* , ranging from 0 to 1. No treatment effect is represented by *t=1* and for 100% effect *t=0*. For modeling chance alone we keep *t = 1*.

For every outcome we take a random *i* within a realistic range. We generate incidence of the outcome within the two groups. For everyone out of the *n* individuals, we generate a random number (*RND*) bet 0 and 1, if *RND < i* the outcome is seen, if *> i* it is not seen. This is repeated independently this for all *n* individuals. The individuals showing the outcome are counted. For the treated group the same is repeated using *RND < i.t* . A chi square test is performed to see whether the difference in the treated and control group is significant, simultaneously the odds ratio (OR) is calculated.

For the next outcome a new random *i* is generated and the process repeated for each of the *o* number of outcomes. At the end, the frequency distribution of total ORs is plotted, indicating those that are significant (figure SI1).

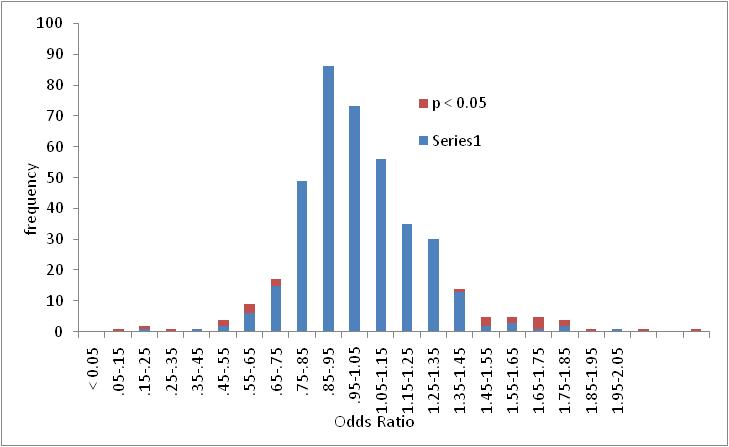

*Figure SI1: A typical frequency distribution of odds ratios (ORs) generated in the absence of any form of dependence. In majority of the distributions the mode lies at 1. However, it can deviate by 10 % (i.e. one bin) or more about 15 % of the times such as in the above. Other parameters for simulation, n =500, 0 = 400, I ranged between 0.01 to 0.2.*

Bringing in dependence in the model:

We assume that there are one or more clusters of outcomes that are interdependent, rest of the outcomes are independent of each other. Dependence is brought in the model by two parameters. *f*= the fraction of outcomes that are completely independent *d* = the number of outcomes in a cluster that are interdependent. So *o(1-f)/d* will be the number of clusters. In a further variation we can randomize the cluster size *d* within a limit for each cluster until all clusters add up to approximately *o(1-f)*.

The simulations first generate data just as in the baseline model for *o.f* number of outcomes. Then one of these outcomes is selected at random. The ratio of the number of cases in the treated group by number in control group is taken as *t* within that cluster i.e. for the next *d* number of outcomes. This is repeated until the total outcomes become *o*.

With 200 to 500 simulation runs for every set of input parameters the resultant distributions of OR, the proportion of outcomes turning out individually significant at p < 0.05 and their position in the distribution is studied.

Results: Introduction of dependence increases the number of false positives, i.e. outcomes that turn out to be individually significant at p < 0.05. While on an average 5 % of the outcomes should turn out to be significant by chance alone, when some degree of dependence is introduced, the average is greater than 5%. In simulations without dependence, the range of outcomes individually significant by chance ranged between 1 and 9. With moderate dependence i.e. *f =0.7* and *d* ranging between 3 and 10, between 4 to 21 outcomes were individually significant. The distribution of significant outcomes was often but not always asymmetrically towards the left or towards the right. Thus even with identical parameters, two runs could have significant heterogeneity or discordance between them.

Violation of independence also causes greater shifts of the mode of the frequency distribution of OR. The probability of the observed shift in the mode of 10 % in the pooled data over all trials was 62 % in simulations using *f = .7* and *d* ranging between 3 and 10. Therefore the observed shift in the mean and median by about 10 % in the clinical data cannot be said to be significant.

The parameters *n, o* and *i* within the range used in the clinical trials being considered. For *f* and *d*, there are no empirical estimates available at present. But simulations show that even for small to moderate level of dependence, i.e. f ranging between 10 and 40 and d between 3 and 10, deviation from baseline results is substantial.

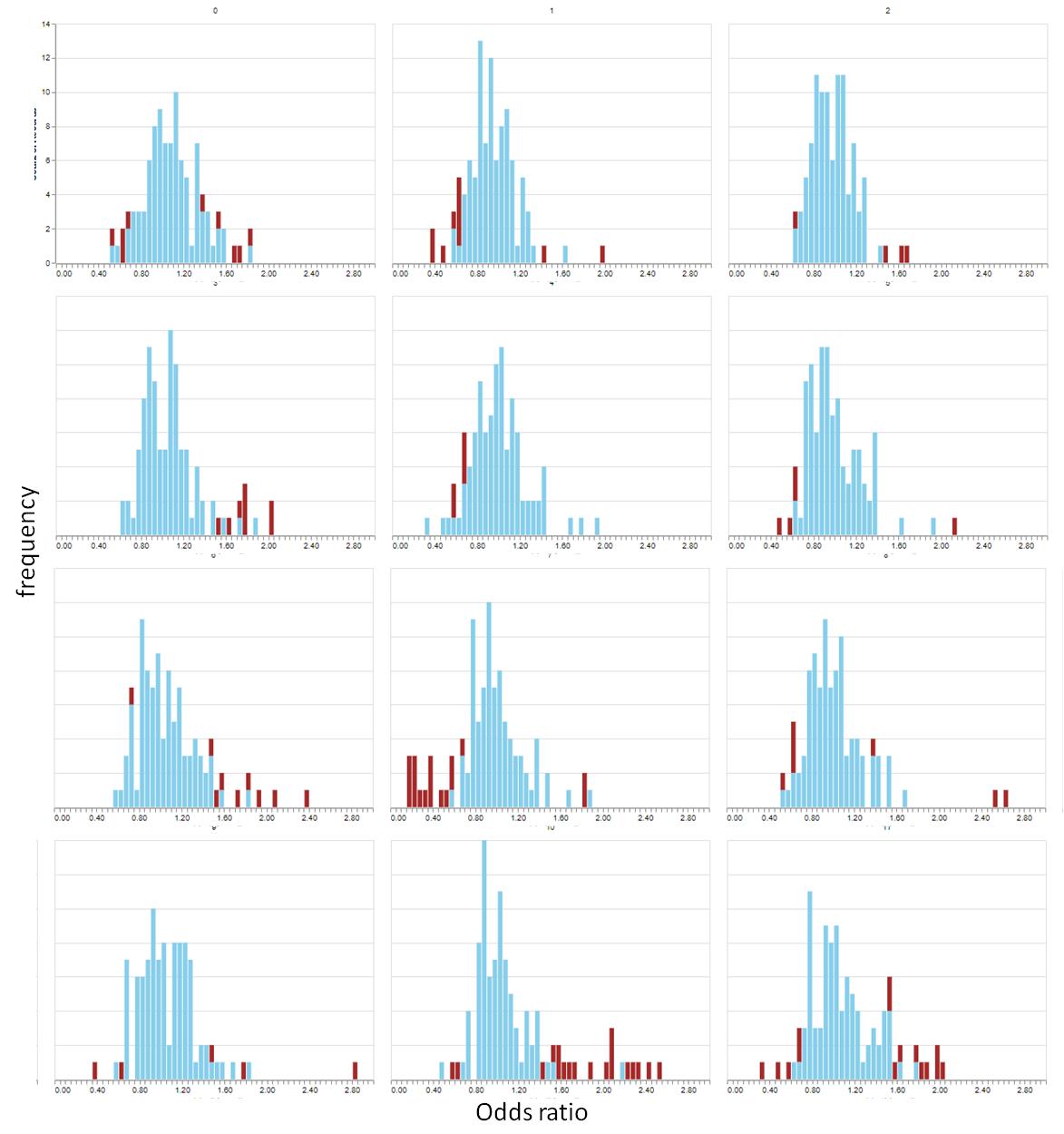

*Figure SI2: The distribution of odds ratios generated by simulation at f = 0.7 and d ranging between 3 and 10. Note that the average number of individually significant results are much greater than the expected 5 %. They are often asymmetrically distributed to the right or to the left. The mode is also commonly shifted from 1.*

The implication of the simulation results for the glucose regulation trials is that, since the assumption of having common pathophysiological pathways is compatible with our current molecular and physiological knowledge, and repetition between aggregate and single end points is obvious, the rate of false positive significance is expected to be considerably greater than 5%. Therefore significance in these trials needs to be interpreted with caution. The total number of outcomes that turned out to be individually significant in the pooled data across the 6 trials is greater than 5 %, (18.7 % including obvious duplications and 8.7 % removing obvious duplications). The 8.7 % are distributed on the two tails nearly symmetrically and this deviation from 5 % is not very large. Therefore the probability of all the significant results of the trials appearing by chance alone is not too small to reject the possibility. The observation that in some of the trials the treatment appeared more beneficial and in other it appeared to cause more harm than good can also be explained by chance alone with a reasonable probability. Since we do not have quantitative empirical estimates of the degree of dependence, we cannot calculate the probabilities exactly. But since the assumption of common pathophysiological pathways is reasonable, serious doubts are raised about the individual significance (at 0.05 level) of some of the outcomes. At present none of the results should be treated as significant.

When we bring in treatment effect, i.e. *t < 1*, the entire distribution shifts to the left. This pattern is highly consistent and the shift in the mode/median increases with decreasing *t*. Owing to its consistency, a shift in mode and median can be considered a more reliable indicator of treatment effect over individual significant outcomes. Simulations suggest that when the hypothesis of a common pathophysiological causal mechanism such as chronic hyperglycemia is being tested using multiple outcomes, a shift in the distribution should serve as a more reliable indicator of treatment effect than individually significant outcomes.
